# Evaluating implementation of LEAPS, a youth-led early childhood care and education intervention in rural Pakistan: protocol for a stepped-wedge cluster randomized trial

**DOI:** 10.1101/2021.04.15.21255571

**Authors:** Aisha K. Yousafzai, Christopher R. Sudfeld, Emily E. Franchett, Saima Siyal, Karima Rehmani, Shelina Bhamani, Quanyi Dai, Chin R. Reyes, Günther Fink, Liliana A. Ponguta

## Abstract

**Background:** The Sustainable Development Goals (SDGs) highlight the importance of investments in early childhood care and education (ECCE) for young children and youth development. Given Pakistan’s large young population, and gender and urban-rural inequalities in access to education, training and employment, such investments offer opportunities. LEAPS is a youth-led ECCE program that trains female youth, 18-24 years, as Community Youth Leaders (CYLs) to deliver high-quality ECCE for children, 3.5-5.5 years, in rural Sindh, Pakistan.

**Methods:** We use a stepped-wedge cluster randomized trial to evaluate implementation of LEAPS. Ninety-nine clusters will be randomized to receive the intervention in one of three seven-month steps (33 clusters/step). Primary outcomes are children’s school readiness (measured with the International Development and Early Learning Assessment) and executive functions. Secondary outcomes are youth personal and professional development, depressive symptoms, and executive functions. Data is collected in cross-sectional surveys of 1,089 children (11 children/cluster from 99 clusters) aged 4.5-5.5 years at four time points (baseline and at the end of each step). We will enroll three youth participant open cohorts, one per step (33 CYLs: 66 comparison youth per cohort; 99:198 in total). Youth cohorts will be assessed at enrollment and every six to seven months thereafter (i.e., once per consecutive Step). A school cohort of 330 LEAPS students (10 students/cluster from 33 clusters) will be enrolled and assessed during Step 1 after intervention rollout and at endline. The quality of the learning environment will be assessed in each LEAPS ECCE center and in a comparison center at two time points midway following rollout and at endline. A concurrent mixed-methods implementation evaluation will assess program fidelity and quality, as well as the extent to which a technical support strategy is successful in strengthening systems for program expansion. A cost evaluation will assess cost-per-beneficiary. Data collection for implementation and cost evaluations will occur in Step 3.

**Discussion:** Youth-led models for ECCE offer a promising approach to support young children and youth; however, there is little empirical evidence on real-world implementation. This study will contribute to the evidence as a means to promote sustainable human development across multiple SDG targets.

**Trial Registration:** ClinicalTrials.gov: NCT03764436. Registered December 5^th^, 2018, https://clinicaltrials.gov/ct2/show/NCT03764436.

## Background

With the ratification of the Sustainable Development Goals (SDGs), the global community embraced a life-course approach to promoting equitable human development for all ages (1). Central to this agenda is the investment in opportunities for lifelong learning and development (2) including Early Childhood Care and Education (ECCE) to bolster early child development and learning outcomes (Target 4.2); teacher-training and workforce development initiatives to enhance quality of educational services (Target 4.c); and youth development programs to promote education, training and employment for youth (Targets 4.4 and 8.6) (1). Innovative, integrated policy and strategies are needed to leverage interconnections across SDG targets in order to promote human development across the lifecourse (3).

An estimated 250 million children under the age of five years in low- and middle-income countries (LMICs) are at risk of not meeting their cognitive developmental potential (4). There is increasing support for ECCE programs as a means to enhance young children’s cognitive, social-emotional, and language development and early literacy and numeracy skills in order to better prepare children to enter primary school (5). A recent meta-analysis found that participation in formal and community-based ECCE programs enhanced cognitive and psychosocial development outcomes among children (6). Although there has been an increase in ECCE programs, inequitable access and low-quality program implementation remains a concern (7). Only 17% of children in LMICs have access to ECCE programs, and disparities in access and quality persist by gender, socioeconomic status, and urbanicity (8). Supporting ECCE program quality is critical in order to benefit young children (6). Investments in ECCE workforce development are essential for maintaining program quality at-scale (8), although questions remain regarding how ECCE programming can be successfully integrated within national systems with fidelity and quality (2).

Lack of access to quality ECCE is also linked to increased school dropout and poor achievement at the primary- and secondary-levels of education (9, 10). In Pakistan, almost 60.7% of preschool-aged children are out of school, and 22.8 million primary and secondary school-aged children are out of school (11). One in six Pakistani children do not transfer from first grade to second grade (12). With many children being out of school or achieving poor learning outcomes, investments in high-quality equitable ECCE programs are needed for girls and boys.

As youth prepare to transition into the labor force, training and employment opportunities are lacking in many settings. Globally, 500 million youth are unemployed, underemployed, or have insecure jobs (13), and youth are nearly three times more likely to be unemployed than adults (14). Approximately 30% of youth aged 15 to 29 years in LMICs are not enrolled in education, employment, or training opportunities (15). These opportunity deficits for youth can in turn lead to longer-term consequences, as youth who are not in employment, education or training upon finishing school are more likely to experience job instability and unemployment and have lower earnings as adults (16). In countries such as Pakistan where 64% of the population is under the age of 29 years (17), programs providing opportunities for youth to pursue education and employment are in high demand. Positive Youth Development (PYD) interventions, defined as interventions that seek to “build skills, assets, and competencies; foster youth agency; build healthy relationships; strengthen the environment; and transform systems” (18), have emerged as a strategy to support youth during this critical phase. While PYD interventions have been well-studied in high-income countries and found to significantly reduce risky behaviors and improve youth mental health, physical health, economic outcomes, and general wellbeing, few youth development programs have been implemented and rigorously evaluated in LMICs (19, 18). A recent systematic review identified just 94 PYD programs targeting youth in LMICs implemented between 1990-2016, of which only 37% of programs had been rigorously evaluated (18). Results indicated that outcome measures were diverse, ranging from measures of impact mediators, to youth behaviors, to youth development (18). Further research is needed to evaluate the effectiveness of youth development interventions in LMICs and to better understand the mediating effects and mechanisms of youth development interventions, particularly among more vulnerable populations such as female youth.

From a life-cycle perspective, investing in young children and youth is critical as both the early childhood and adolescent years are sensitive periods of brain development. The early years of a child’s life are the first window of opportunity to positively contribute to children’s development, establishing a strong foundation for lifelong health and learning (20). Late adolescence and youth is a second period when the high plasticity of the brain leads to significant intellectual and social-emotional maturity, referred to as the second window of opportunity (21, 22). During both of these developmental periods, executive functioning skills are undergoing a phase of rapid development. Executive functions are the skills that support emotional regulation, social skills, attention, and reasoning skills and are essential to children’s and youth’s successful performance in school, work and in fostering healthy relationships (23, 24). As children’s executive functioning skills are thought to develop through interactions with adults possessing strong executive functions (25), supporting executive function development among caregivers may bolster gains in these skills among children (26). Engaging youth as ECCE providers offers an innovative solution to enhance executive functioning skills and increase opportunities for both young children and youth. A recently published conceptual model for youth-led ECCE outlines a strategy to promote both child and youth development, by providing appropriate training and mentoring for youth to enable them to deliver high-quality ECCE programming (27). While examples of youth-led ECCE programs are limited (27), a few key examples can be found in LMICs, including the Gandhi Fellowship program in western India (28) and the Early Childhood Development (ECD)-practitioner training track of the Economic Empowerment of Adolescent Girls and Young Women (EPAG) project in Liberia (29, 30). Youth-led ECCE offers a promising model for implementing a lifecourse approach to human development.

### Evidence for Youth-led ECCE from Pakistan

*Youth <u>L</u>eaders for <u>E</u>arly Childhood <u>A</u>ssuring Children are <u>P</u>repared for <u>S</u>chool* (LEAPS) is a youth-led ECCE program which was conceptualized as a response to the education and training needs of children and youth in rural Sindh, Pakistan. LEAPS was developed following formative research to identify local barriers and enablers of ECCE and PYD interventions. LEAPS trains female youth aged 18-24 years with a minimum 10th grade education as Community Youth Leaders (CYLs), who serve as local advocates for ECD and deliver quality ECCE programming for children aged 3.5-5.5 years in community-based preschools. A pilot of LEAPS was implemented in partnership with the Government of Pakistan’s National Commission for Human Development (NCHD) in 2016 in district Naushahro Feroze (26). As proof-of-concept, LEAPS was evaluated through a small-scale cluster-randomized controlled efficacy trial. Five villages were randomized per arm, and a total of 240 children aged 3.5-5.5 years were enrolled (170 children per study arm). Intervention clusters received the LEAPS program, and control clusters received education services as usual. The results showed significant improvements in children’s school readiness as assessed by the International Development and Early Learning Assessment (IDELA; 31) (Cohen’s d = 0.3) (26). A qualitative analysis of CYL exit interviews also indicated improved professional and personal development benefits for female youth leaders including aspirations for education and career, mental health benefits, and higher self-confidence (32, 33).

### Objectives

The present study has five objectives:

- Objective 1: To evaluate the impact of the larger-scale implementation of the LEAPS model on children’s school readiness and executive functioning;
- Objective 2: To evaluate the impact of the larger-scale implementation of the LEAPS model on female youth development;
- Objective 3: To evaluate the larger-scale implementation of the LEAPS model as integrated in NCHD platforms and delivered through NCHD systems in order to assess fidelity, demand, enablers and barriers to implementation and quality;
- Objective 4: To evaluate the readiness of NCHD to uptake LEAPS for sustainable-scaling and replication in other districts and provinces in Pakistan, with respect to leadership and governance, training, supervision, advocacy and communication, monitoring and evaluation, and financing and resource mobilization; and
- Objective 5: To examine cost-per-beneficiary ratios through a cost evaluation.

It is hypothesized that after implementation of LEAPS, clusters will experience significantly improved population-level outcomes for school readiness among children aged 4.5-5.5 years as assessed by the International Development and Early Learning Assessment (IDELA), as well as improvements in children’s executive functioning skills. We also hypothesize that female youth who participate in LEAPS will have improved outcomes for executive functioning, and personal and professional development relative to youth who are not LEAPS participants.

## Methods

Methods are described according to the SPIRIT Guidelines (34).

### Study Setting

The current trial will be conducted in rural Sindh, Pakistan, across four districts: Naushahro Feroze, Khairpur, Sukkur, and Dadu. The ECCE participation in this region is low. A recent report indicated that 54% of children aged three to five years in rural Sindh were not enrolled in any ECCE program (11). The lack of school readiness is also reflected in child learning levels in Grade 1 with less than half of children surveyed able to identify Sindhi letters or English letters, or to recognize numbers one to 99. Low literacy rates among parents present additional challenges with three-quarters of mothers and over half of fathers without primary school completion (Grades 1-5), which can limit parents’ support for their child’s at-home learning (11) (see Table 1 in Appendix 1). Communities in rural Sindh also face significant gender disparities in school enrollment and literacy rates among youth: the literacy rate for male youth (61%) is more than twice that of female youth (30%) (35). Trends in Pakistan more broadly indicate that female youth are also at a significant disadvantage in employment (36) and thus require more targeted support in education, training, and work (see Table 2 in Appendix 1).

**Table 1.**
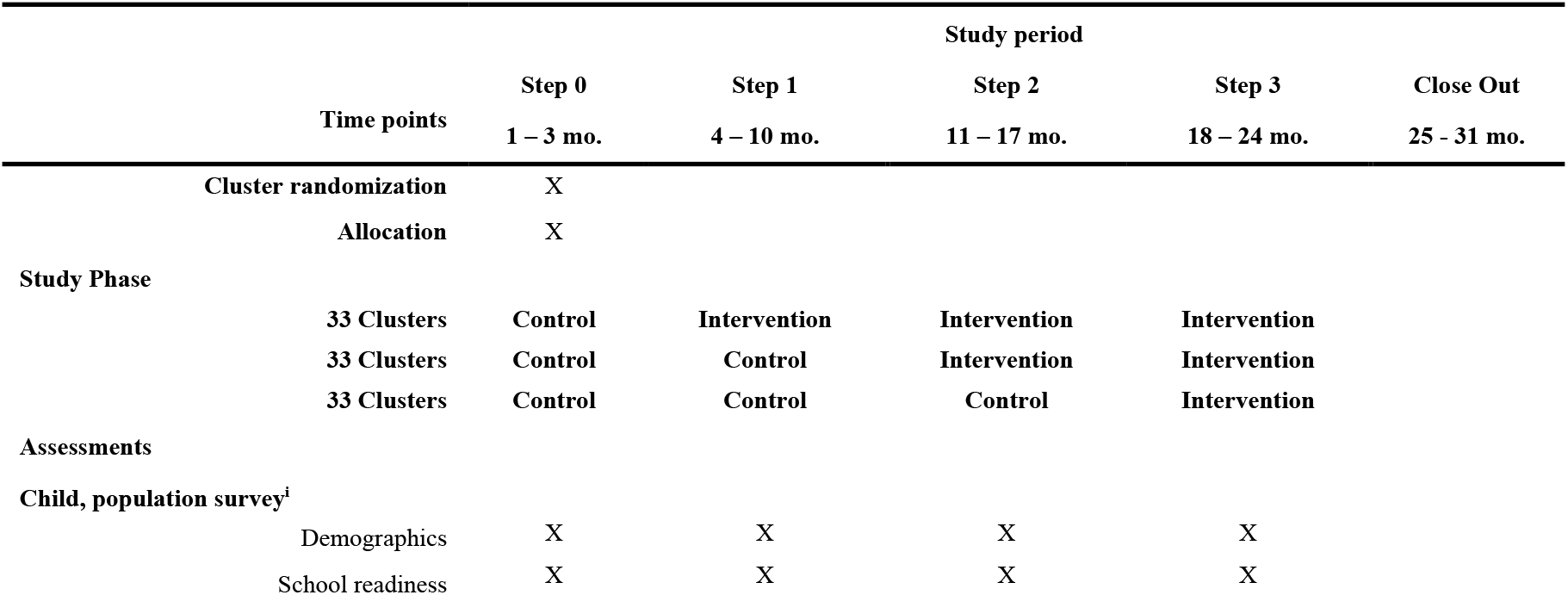
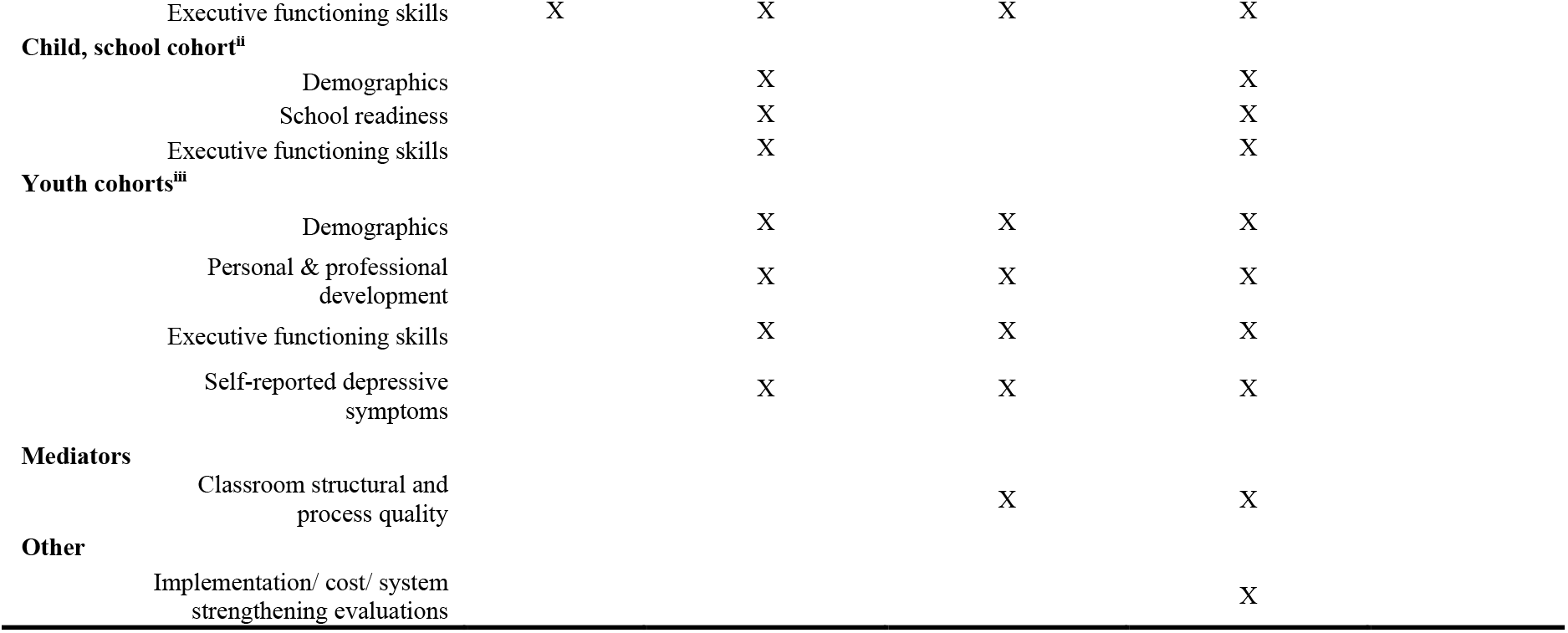
SPIRIT figure depicting schedule of enrollment, interventions, and assessments. iThe population survey is a cross-sectional survey enrolling 11 children aged 4.5-5.5 years and their primary caregiver per cluster in each of the 99 clusters (total of 1,089 child-caregiver dyads per round for four rounds). iiThe school cohort is a closed cohort enrolling 10 LEAPS preschoolers and their primary caregiver per cluster (stratified by gender) in each of the 33 clusters receiving the intervention during Step 1 (total of 330 child-caregiver dyads). iiiAn open cohort of youth participants will be enrolled in each of the Steps (3 cohorts in total). Each cohort includes all Community Youth Leaders (CYLs) hired by NCHD, as well as two comparison youth participants per cluster (33 CYLs and 66 comparison youth per cohort; 99 CYLs and 198 comparison youth in total).

**Table 2.** Qualitative Data Collection Activities and Participants. Table 2 summarizes planned qualitative data collection activities by stakeholder, including sampling details. FGD = Focus Group Discussion, IDI = In-depth Interview.

| Stakeholder | No. of IDIs/ FGDs | Participant Sampling |
| --- | --- | --- |
| CYLs | 4 FGDs | • Representation from all districts and Steps<br>• Maximize variation in CYL age, marital status, education, parenting status |
| CYLs | 16 -17 IDIs | • Representation from all districts and Steps<br>• Variation in marital status, parenting status, age, and urbanicity |
| CYLs' Families | 4 Family Interviews | • Sub-set Families of the same CYLs in IDIs<br>• Variation in marital status and family structure |
| Children's Mothers | 4 FGDs | • Representation from all districts<br>• Child must be enrolled in LEAPS preschool<br>• From rural communities |
| Children's Fathers | 4 FGDS | • Representation from all districts<br>• Child must be enrolled in LEAPS preschool<br>• From rural communities |
| NCHD Feeder school teachers | 4 FGDs | • Representation from all districts and Steps |
| NCHD Field Officers | 4 FGDs | • Representation from all districts and Steps<br>• All female Field Officers will be enrolled<br>• Variation in number of supervisees |
| NCHD Master Trainers | 1 FGD | • Including all Field Officers who have been selected as Master Trainers |
| LIST Team | 1 FGD | • Including the LIST members |
| LIST Members | 1 FGD (Pile sorting activity) | • Including the LIST members |
| NCHD District team Managers | 1-2 FGDs | • Including all NCHD district managers |
| District Stakeholders | 4-8 IDIs | Sample most relevant/influential stakeholders per district as per Intervention Team's inputs:<br>• Village leaders<br>• District government officials<br>• Social Welfare Department<br>• Education Department |

### Implementation partner and LEAPS intervention support team

Pakistan’s National Commission for Human Development (NCHD) was identified as the partner for program design and implementation during the formative research phase. The NCHD is an autonomous federal body established under the office of the President in 2002 mandated to support and augment human development efforts in Pakistan. NCHD works in conjunction with the government’s health and education sectors and with local communities to fill service implementation gaps. Currently NCHD is present in 124 districts of Pakistan (37). NCHD’s services include provision of primary education for children, vocational training programs for youth, and health services for communities in the most remote areas of Pakistan. The NCHD’s Universal Primary Education (UPE) program trains teachers who are then hired to teach in “Feeder” primary schools (Grades 1-5), located in remote areas where government schools are not operating. The NCHD has also previously provided vocational training for young women, including tailoring courses and the provision of sewing machines.

The current study engages NCHD leadership at the Federal Office, Sindh Province Office, and four district offices, as well as the district-level NCHD Field Officers (FOs), who will be responsible for supervising the CYLs in their catchment area. All FOs have a minimum bachelor’s-level education and several years of experience working with NCHD’s Feeder primary school teachers and local community. Implementation of LEAPS activities will be led by NCHD, with technical support from the LEAPS Intervention Support Team (LIST), a five-member female team, all of whom are local to the implementation communities, with a Master’s-level education and several years of experience delivering community interventions including ECCE. The purpose of the LIST group is to provide training and technical support for NCHD to promote the sustainable rollout and integration of LEAPS within NCHD’s UPE program. All LIST members will receive a three-month training prior to rollout of LEAPS, followed by a three-day refresher training and ongoing on-the-job mentorship and support in coaching, supervision, and coordination from the LEAPS research team.

### Description of the LEAPS intervention

The details of the intervention are reported in accordance with the TiDiERS guidelines (38). The Implementation Map (Figure 1) details the program delivery systems and anticipated outputs, outcomes, and impact. A summary of the dosage, location, modes of delivery, and monitoring tools for key LEAPS intervention activities is presented in Table 3 of Appendix 1.

**Table 3.**
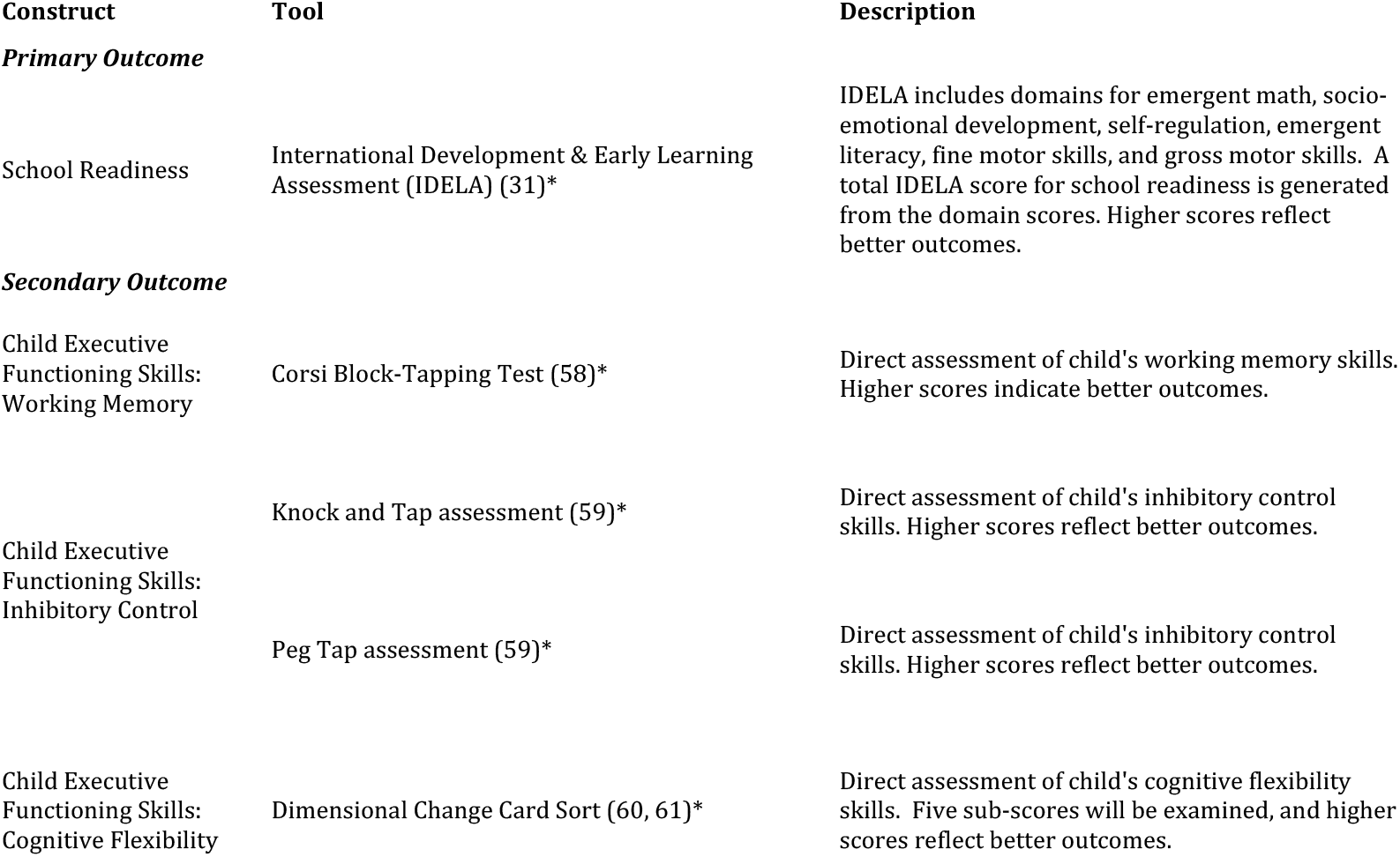
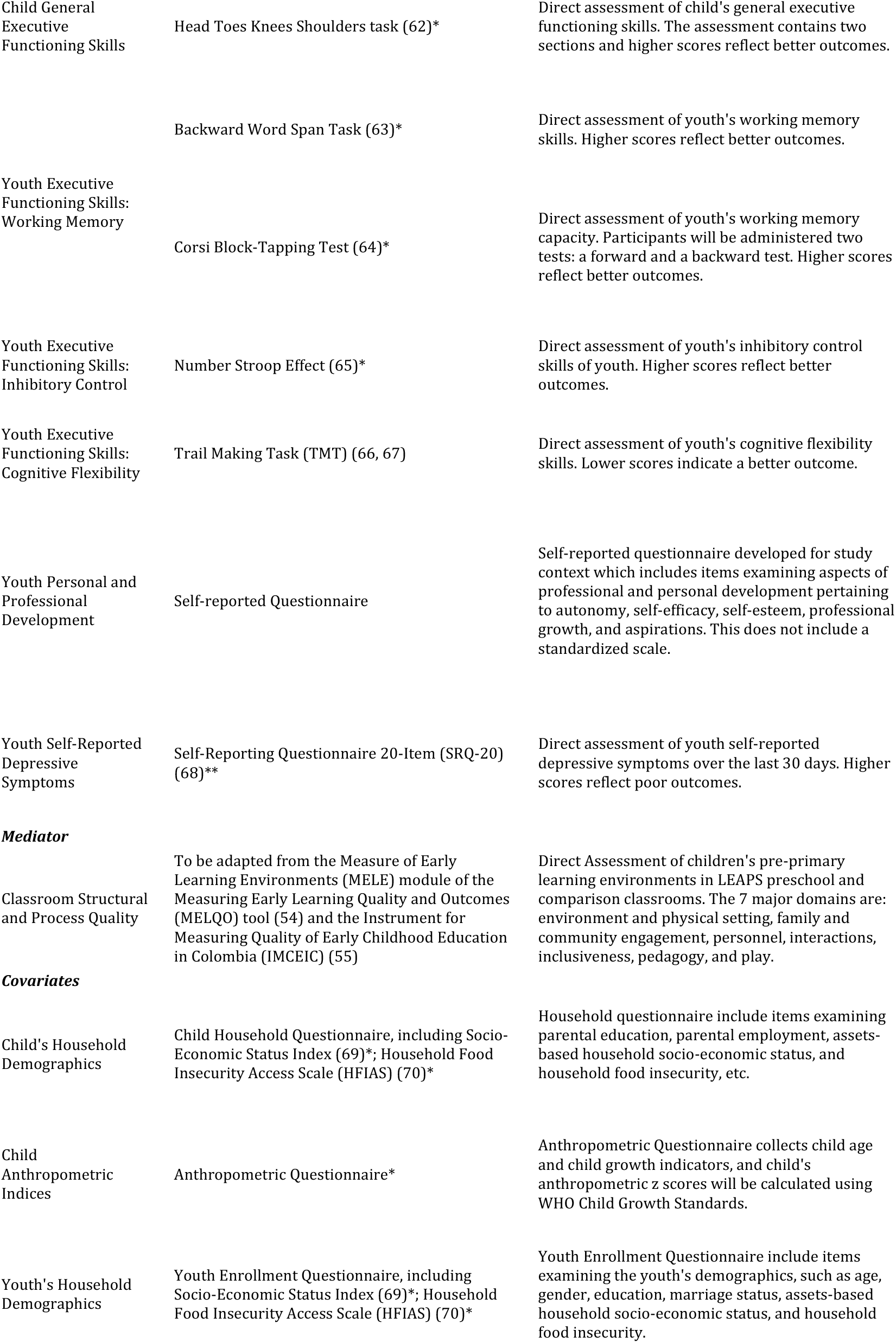
Description of Trial Outcomes, Assessment Tools, Mediators, and Covariates. *The starred IDELA, Executive Functioning Skills assessments, anthropometric, and demographic measures have previously been adapted for the rural Pakistan context for use in the LEAPS efficacy trial (26). **The SRQ-20 was previously adapted and validated for use in rural Pakistan(57). All other measures were adapted or developed for the use of this trial.

**Figure 1.**
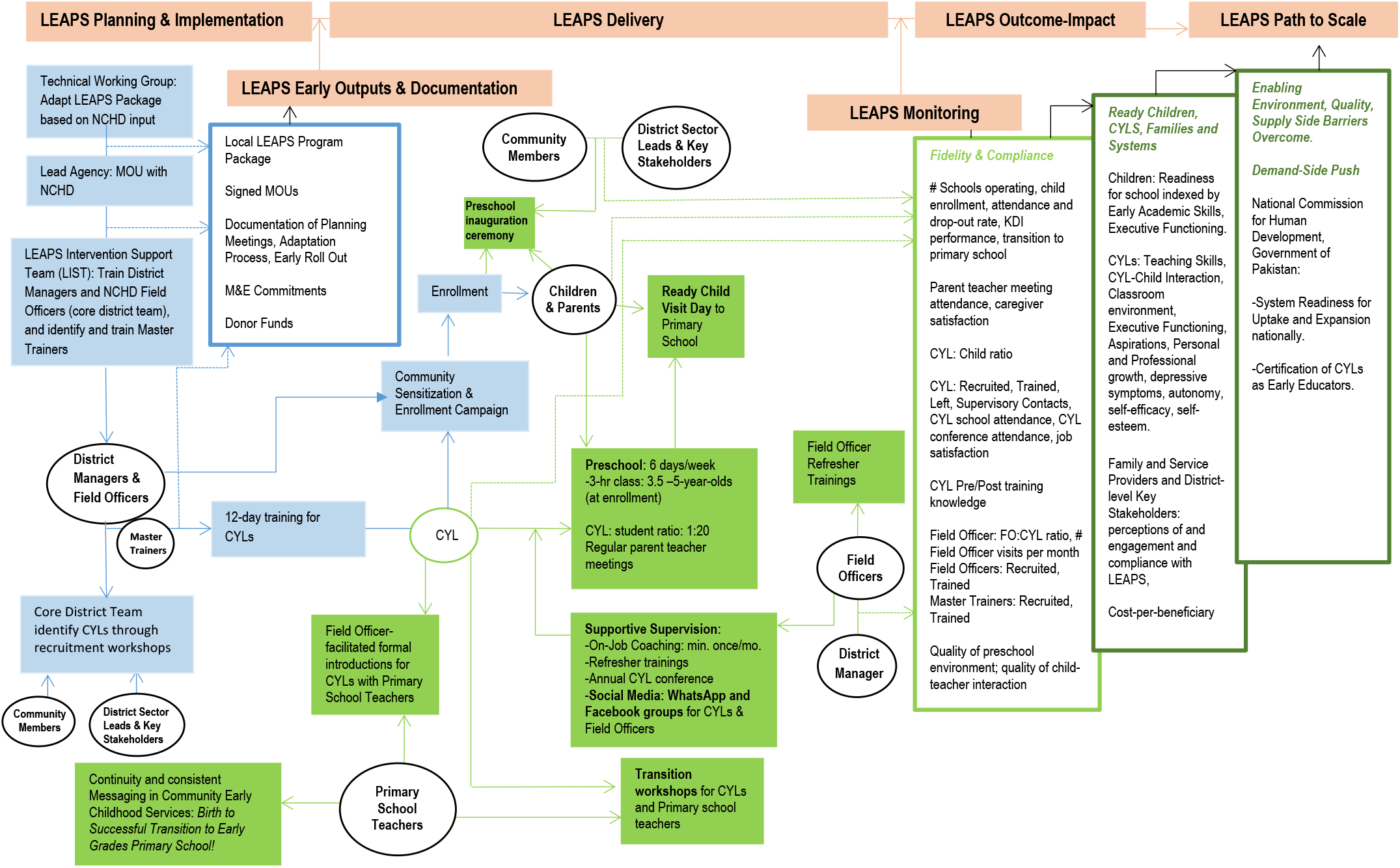
LEAPS Implementation Map presenting the intervention delivery systems. The solid blue figures represent the early program inputs. Solid green figures represent the core program components. The three boxes on the far right describe intended outputs, outcomes, and eventual impact and sustainability at scale. CYL=Community Youth Leader, M&E=Monitoring and evaluation, MOU=Memorandum of Understanding; NCHD= National Commission for Human Development.

#### Training of NCHD Field Officers and Master Trainers

An initial five-day Training of Trainers (TOT) will be held for the 42 FOs and eight district managers who will be engaged in LEAPS. The TOT will be implemented by the LIST members using a training manual. The training introduces basic concepts of ECCE, familiarizes participants with the LEAPS program model, and targets essential supervision and coaching skills, to prepare FOs for their role as CYL supervisors. The LIST members will use an observation checklist to assess FOs’ performance during the training. After the training, LIST members and district managers will jointly select the ten highest-performing FOs to be the LEAPS Master Trainers (MTs). In addition to their supervision responsibilities, the MTs will lead all CYL training activities in their district. The MTs will receive training per diems aligned with the government per diem rates for their employee grade level. LIST members will provide a one-day preparatory training for MTs prior to each CYL training, along with coaching and feedback during training sessions. LIST members will additionally provide ongoing support for all FOs during weekly group coordination meetings at district offices, periodic group refresher trainings providing targeted support for specific skills, and monthly one-on-one field visits to provide on-the-job coaching during CYL supervision.

#### CYL recruitment

Following the TOT, district managers and FOs will initiate the CYL recruitment process in their catchment areas. NCHD will engage Feeder school teachers, community health workers, village leaders, and other community members to inquire about potential female youth candidates meeting the CYL recruitment criteria (i.e., aged 18 to 24 years old, have completed Grade 10 schooling, have their family’s support to participate). Candidates will be invited to attend a local CYL recruitment workshop. Recruitment workshops will be led by district managers and FOs, with support from LIST members, and comprise a range of activities designed to assess candidates’ literacy, numeracy, general knowledge, attitudes towards working with young children, problem solving, creativity, communication and leadership potential. Youth participants’ performance on workshop activities will be scored using a standardized rubric. Participants who meet the CYL recruitment criteria and have the highest scores will be selected as the CYL.

#### CYL training

Selected CYLs will be invited to attend their district’s 12-day teacher training. Training will introduce basic concepts of early childhood development and ECCE, orient CYLs to the LEAPS curriculum, and provide hands-on opportunities to practice activities from the curriculum (see Table 4 in Appendix 1 for training overview). We intend to modify consecutive trainings based on lessons-learned from early training activities. The CYLs will continue to receive on-the-job vocational training through supportive supervision visits from FOs and LIST members, as well as through participation in CYL refresher trainings and other youth development activities, including an annual CYL conference. Once hired, CYLs will receive a monthly salary of Rs. 8000 (approximately $50 USD), which aligns with the NCHD salary guidelines for teachers. CYLs will additionally receive travel support for all training days and professional development activities.

#### Preschool space identification and setup

NCHD district offices will be tasked with identifying a space for the LEAPS preschool in each cluster through a community engagement strategy. To be considered viable, a preschool space must have a roof, enclosed walls, access to a washroom, access to a water source, and a door that locks. The preschool space will be an in-kind donation from the local community. The CYLs will each receive a LEAPS classroom starter-kit, which includes learning materials, furniture, and first aid and cleaning supplies. The CYLs and community members will collaborate to set up the preschool space, keeping in mind the elements for a high-quality preschool program that are relayed during trainings.

#### Preschool enrollment drives

After the preschool is set up, CYLs will conduct enrollment drives in the village. Children will be eligible for enrolment if they are aged 3.5-5 years and reside locally. CYLs will be coached to ensure equal opportunities for both girls and boys to enroll in LEAPS. Target enrollment will be 20 children per classroom. CYLs will place any additional children on a waiting list.

#### Inauguration ceremony

Prior to opening the LEAPS preschool, the FO and CYL will plan an inauguration ceremony to celebrate with parents of LEAPS children, local leaders, and the wider community. Inauguration ceremonies offer an essential opportunity to further engage with the community and raise awareness for LEAPs and the importance of ECCE. Following the inauguration ceremonies, preschool classes will officially commence.

#### Preschool classes and curriculum

LEAPS preschools will follow the NCHD UPE program academic calendar and six-day school week. LEAPS school days are three hours long and follow a structured classroom routine (see Table 5 in Appendix 1). The LEAPS curriculum was developed through formative research, drawing on the principles of the HighScope preschool curriculum and aligned with the Government of Pakistan’s early childhood education curriculum. It is designed to support the achievement of 50 Key Development Indicators (KDIs) across seven learning areas (presented in Table 4 of Appendix 1). CYLs will record children’s attendance daily. CYLs will use a Child Progress Form to track children’s progress toward achieving the 50 KDIs. CYLs will be coached to monitor the overall class performance and to share general strengths and areas for additional support with parents during Parent Teacher Meetings (PTMs). PTMs will provide an opportunity for CYLs to teach parents more about the LEAPS program and their child’s learning needs.

#### Transition from LEAPS to primary school

In alignment with NCHD’s Feeder school enrollment criteria, at the end of the academic year, LEAPS children who are aged five years and above will transition to the NCHD Feeder school. The NCHD may recommend that children who are between ages 5 – 5.5 years who have not yet achieved their 50 KDIs continue to attend LEAPS to boost their KDI achievement prior to transitioning to primary school at age 5.5 years. A Transition Workshop will be conducted for all CYLs and NCHD Feeder school teachers with the goal of preparing the Feeder school teachers to welcome and support the LEAPS graduates. Additionally, the CYLs will accompany LEAPS children to visit their Feeder school to familiarize students with their new teacher, school environment and routine.

#### Supervision visits

In order to ensure CYLs are supported to offer high-quality preschool programming to children, CYLs will receive monthly on-the-job supervision visits from their FO. Visits will last for the entire preschool session. The LIST members will in turn provide ongoing support and coaching to FOs by accompanying them on their preschool supervision visits once a month. The NCHD FOs use a supervision checklist tool during their visits to assess program quality and fidelity, and to provide constructive feedback to the CYL on her strengths and areas for improvement. Similarly, the LIST member uses a separate supervision checklist tool to observe and provide constructive feedback to the FO.

#### Refresher trainings for CYLs and FOs

Both CYLs and FOs will participate in one-day professional development workshops or refresher trainings. For CYLs, these will be led by NCHD MTs, and for FOs these will be led by LIST members.

1. Core refresher trainings:
  a. CYL refresher trainings: A set of core training topics including preschool routine, child progress forms, and preschool behavior management will be offered to CYLs to reinforce key ECCE skills. These are intended to be held quarterly.
  b. FO refresher trainings: A set of training topics including use of the supervisory checklist tool, child progress forms, and gender and youth-sensitive supervision are offered to FOs to ensure high-quality preschool supervision visits. These are intended to be held bi-annually.
2. Needs-based training: Additional professional development workshops may be held to address observed needs of CYLs and FOs identified by NCHD and LIST members.

#### Annual CYL Conference

An annual conference will be held for all CYLs in one of the four targeted districts. The conference will provide CYLs with a platform to meet their peers, share ideas, discuss career-related challenges and ideas, and brainstorm solutions. The purpose of the conference is three-fold: (i) to promote female youth agency; (ii) to support female youth personal and professional development; and (iii) to showcase the CYLs’ work.

#### LEAPS Response During Emergencies and Conflict

In the past there have been cases of political unrest and major flooding in Pakistan, which has led schools to shut down for weeks (39). Table 6 in Appendix 1 details the actions that LEAPS will undertake to support CYLs and LEAPS children in the event of emergency response.

### Study design

This transition-to-scale trial uses a stepped-wedge cluster-randomized design with three Steps to evaluate the impact of the LEAPS program on children’s school readiness and female youth development in 99 clusters. A cluster-randomized approach was selected as the point of intervention will be at the community-level; therefore, this approach minimizes the potential for contamination between intervention and non-intervention recipients. A stepped-wedge approach was selected as a pragmatic approach to support program implementation on a larger scale (40). The LEAPS program will be introduced in clusters in a randomized sequence at three discrete time points over the course of the 31-month trial. At each of the three time points, 33 clusters will cross over from the control condition (i.e., NCHD services as usual) to the intervention condition (i.e., initiating the LEAPS program) (see Figure 2). Once a cluster crosses over it will remain exposed (i.e., the program will remain in operation) for the duration of the trial. Each Step will be approximately seven months in duration. Prior to opening the LEAPS schools in each Step, targeted clusters will undergo a transition period in which NCHD, with technical assistance from the LIST members, will initiate CYL recruitment and trainings, preschool space identification, and community sensitization activities.

**Figure 2.**
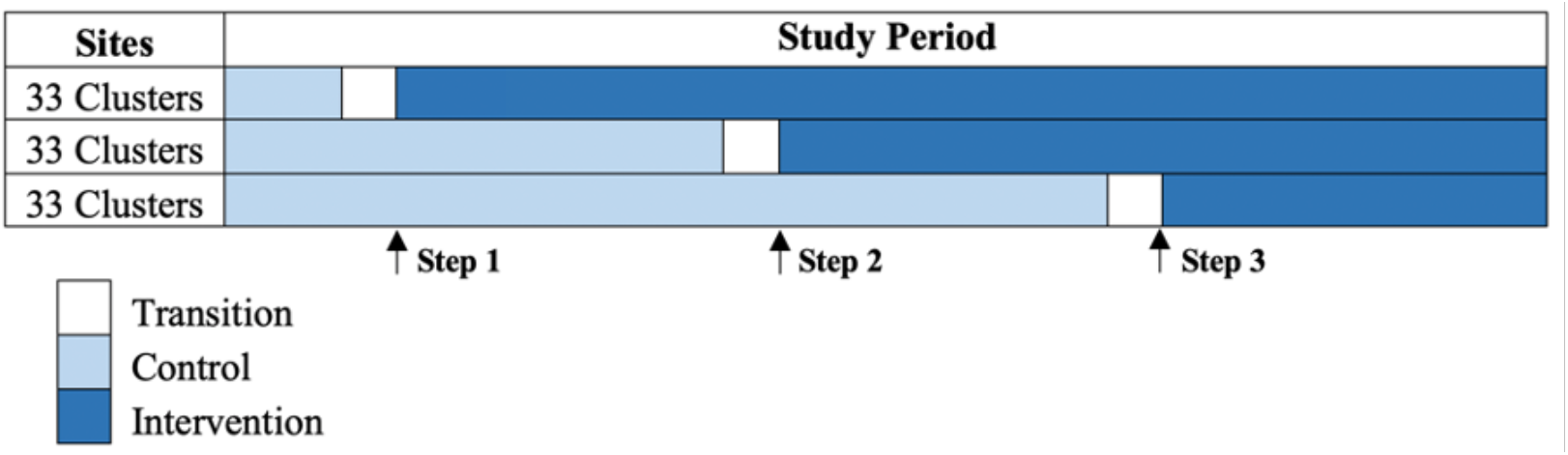
Stepped-Wedge Cluster Randomized Trial Timeline

### Cluster Identification & Randomization

A cluster is defined as the catchment area of an NCHD Feeder primary school. We restricted the study to NCHD Feeder school catchment areas because the possibility to transition from ECCE to primary school was considered essential. The research team coordinated with the NCHD to compile a list of all villages across the four districts with NCHD Feeder schools (n=119), excluding the villages from the LEAPS pilot trial. With the NCHD, the research team screened potential clusters against predefined exclusion criteria, including (i) lack of an eligible CYL candidate, (ii) safety and security concerns, (iii) lack of proper space for the preschool, or (iv) NCHD Feeder school had closed. Twenty clusters were excluded though this process. We applied stratified randomization to the 99 clusters using the 4 districts as strata with 3 blocks in each stratum. The 99 clusters were randomly assigned in blocks to one of the three Steps (n= 33 clusters per Step). The stratified randomization was completed using a computer-generated random allocation sequence in Excel. Block randomization was used to ensure that clusters were evenly dispersed by geography across each Step, so that intervention rollout was feasible for implementation partners. The co-investigator, and the project Senior Statistician (CS), who conducted the randomization is not involved in any of the field activities. The final trial includes six clusters in Dadu, nine clusters in Sukkur, 28 clusters in Khairpur, and 56 clusters in Naushahro Feroze (Figure 3).

**Figure 3.**
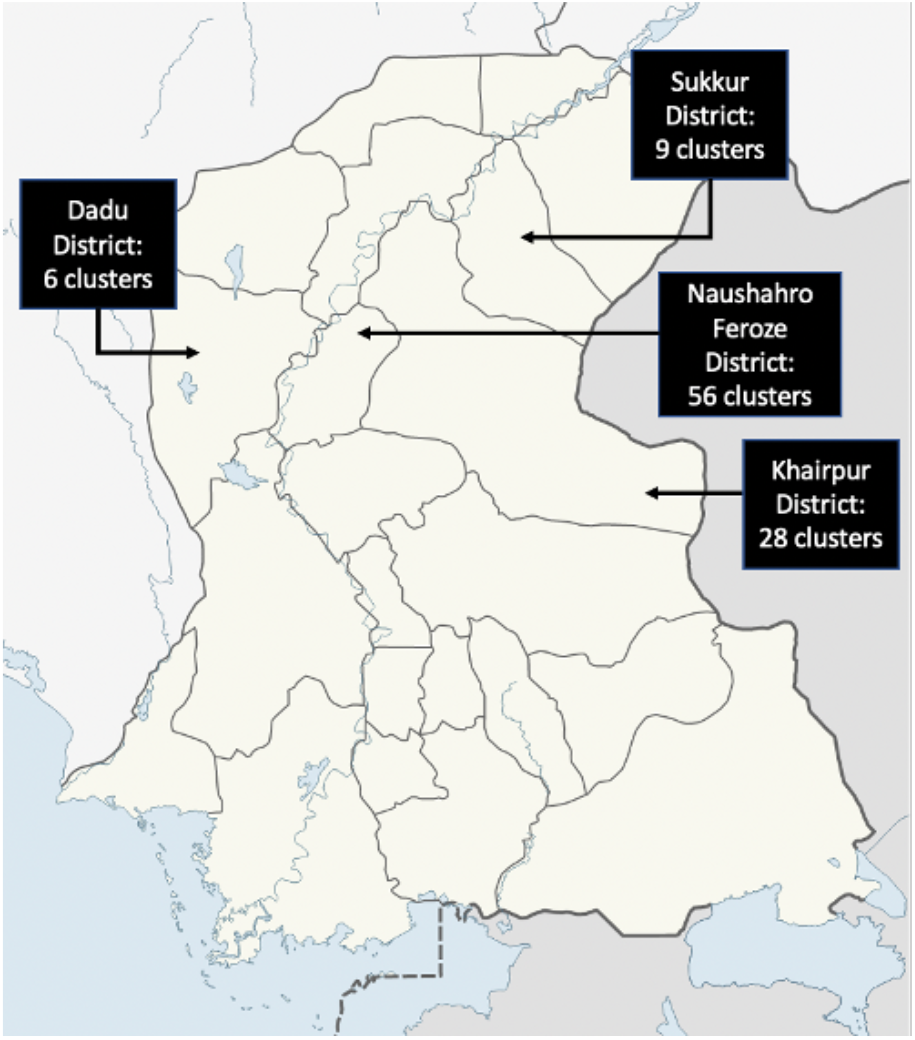
Map of Districts for the LEAPS Transition-to-Scale Trial in Rural Sindh *Note: One cluster from district Khairpur was randomized with district Naushahro Feroze clusters, both because it was nearer in terms of location and a multiple of 3 was needed in each group*.

### Description of data collection activities & study sample

Data collection activities and the participant timeline are summarized in the SPIRIT figure (Table 1) (34). The trial will include the following data collection activities:

#### Population survey

A cross-sectional survey of child-caregiver dyads will be conducted at four time points: at baseline (Step 0; mos. 1-3) and again at the end of each consecutive Step, with six to nine months between data collection rounds. Eleven children aged 4.5-5.5 years and their caregivers will be enrolled from each of the 99 clusters (1,089 child-caregiver dyads total), regardless of the child’s enrollment in LEAPS. The primary objective of the population survey will be to assess changes in the average school-readiness over time. By including both LEAPS participants and children who are not participating in the LEAPS program, this design will allow us to separate treatment effects on the treated from average (intent-to-treat) effects at the population level. The sample will be drawn from village birth registries provided by NCHD, which will be verified a) by cross-checking against birth registries from local community health workers; and b) through household visits conducted by research team members. The sample will be re-drawn at each of the four data collection time points. Children may be enrolled for assessment at multiple time points. Assessors will use a random number generator to select 11 participants/cluster using the verified lists of eligible children prepared for each cluster. If a participant refuses, assessors will select the next participant in the random sequence.

- Eligibility criteria: Child is age 4.5-5.5 years +/- 2 weeks and resides in the targeted village
- Exclusion criteria: Child or caregiver shows signs of severe clinical health condition or disability

#### School cohort

A closed cohort of 330 LEAPS children and their caregivers (10 children/cluster, stratified by gender) will be enrolled from the 33 clusters receiving the intervention in Step 1. The main purpose of the school cohort is to assess individual learning and growth trajectories among children participating in the program. Child-caregiver dyads will be enrolled and assessed during Step 1, after the intervention has been initiated in month eight of the trial and followed up again at the end of Step 3 in month 24. Assessors will use a random number generator to randomly select five girls and five boys from the CYL’s enrolment register. If a participant refuses, the next participant in the random sequence will be invited. If fewer than five participants of one gender are registered in the LEAPS class, additional children of the other gender may be enrolled to meet the ten-student sample for that cluster.

- Eligibility criteria: Child must be registered and regularly attending the LEAPS school
- Exclusion criteria: Child or caregiver shows signs of severe clinical health condition or disability

#### Youth participants

A case:comparison design will be employed in order to examine program impacts on youth participants. In each Step, we will enroll an open cohort comprising the 33 CYLs from the 33 clusters introducing the intervention in that Step (one CYL/cluster), as well as 66 youth comparisons for a 1:2 CYL: youth comparison ratio (i.e., 3 youth cohorts (one/Step) enrolling 33 CYLs:66 youth comparisons; in total, 99 CYLs and 198 youth comparisons enrolled). In each Step, the CYLs will be enrolled in the evaluation and assessed as they are hired by NCHD, during the month before and first month of the Step (i.e., in mos. 3-4 for Step 1, in mos. 10-11 for Step 2, and in mos. 17-18 for Step 3). The youth comparisons will be enrolled and assessed beginning the month before through the first two months of the Step (i.e., in mos. 3-5 for Step 1, in mos. 10-12 for Step 2, and in mos. 17-19 for Step 3). Youth cohort participants will be followed up every six months. If a CYL drops out of the LEAPS program, she will be invited to remain in the evaluation as a comparison youth. Similarly, if NCHD hires a comparison youth as a CYL, she will be invited to continue in the study as a CYL, and a new comparison youth will be identified and enrolled.

Youth comparisons will be identified using a screening tool during CYL recruitment activities and through door-to-door household visits. The screening tool for youth comparisons includes a series of questions and short case study exercises to ensure that the participant, i) meets the same gender, age and education requirements as the CYL, ii) has support from her family to pursue employment outside of the workplace (as is the case with CYLs), and iii) demonstrates confidence, and advocacy and problem-solving skills. The tool takes approximately 15 minutes to implement. These screening criteria represent the qualities that the research team considers most central to the CYL role. Given that CYLs may be positive deviants in their community and have demonstrated high-selection workshop performance relative to their peers, the purpose of the screening tool is to ensure to the best of our ability that youth participants are a fair comparison for CYLs in terms of family environment and skillsets baseline.

- Eligibility criteria:
  - CYLs must be hired by NCHD.
  - Youth comparisons must be female, aged 18-24 years +/- 2 weeks, have a minimum tenth grade education, have support from family members to pursue employment, and meet capacity-checks for confidence, advocacy and problem-solving skills.
- Exclusion criteria: Youth participant shows signs of severe clinical health condition or disability

#### Classroom Observations

To examine quality of the preschool environment, classroom observations will be conducted in both LEAPS classrooms and comparison preschool classrooms. As LEAPS schools are by design typically the only preschool service operating in an intervention cluster, in order to select the most suitable comparison, the comparison preschool classrooms will be selected based on: i) location: school should be located as close to the LEAPS evaluation cluster as possible; and ii) prioritizing enrollment of government schools (standard service) where available, followed by NCHD-operated schools and other options (private/Madrasa) to reflect the distribution of these types of schools in rural communities in the targeted districts. Comparison classrooms may vary with respect to the preschool classroom arrangements, including grade-level groupings (stand-alone preschool classrooms or multi-grade classrooms), single- or mixed-gender classroom, and informal or formal settings. One comparison classroom teacher will be enrolled for each LEAPS school. All Step 1 and Step 2 LEAPS schools (n=66) and equivalent number of comparison schools will be observed at two time points: once in Step 2 (mos. 14-16) and once during Step 3 (mos. 21-24). All Step 3 LEAPS schools (n=33) and equivalent number of comparison schools will be observed at one timepoint in Step 3 (mos. 21-24).

Eligibility criteria:

∘ LEAPS school: Must be located in an intervention cluster
∘ Comparison preschools: Fits criteria for location, type of school, and gender make-up
• Exclusion criteria: None

#### Implementation, costing and systems strengthening evaluations

A phenomenological design will be employed to collect original qualitative data in addition to administrative and programmatic data, Global Positioning System (GPS) and location data, and costing data to inform the concurrent mixed-methods implementation evaluation, cost evaluation, and systems strengthening evaluation (all three of which are described in detail in the sub-sections below). Data for these activities will be collected in mos. 22 to 24 at the end of Step 3. Participants in qualitative data collection activities will be purposefully sampled to reflect variation in district, demographics, and stakeholder role. Qualitative data collection will include focus group discussions (FGDs) and in-depth interviews (IDIs). Administrative, classroom and programmatic data will be collected from NCHD district offices and around training and supervision activities. Costing data will be collected from internal project budgets (NCHD program and university research partners).

- Eligibility Criteria: Sampling and participant details for qualitative data collection are detailed in Table 2
- Exclusion Criteria: Participants demonstrate signs of severe clinical health condition or disability

#### Implementation evaluation

Implementation evaluations are critical in transition-to-scale research for a number of reasons: i) implementation evaluations provide key insights on the challenges of scaling up, in terms of replicability, coverage, appropriate monitoring and evaluation and sustainability (39); ii) they help to explain why certain results were achieved, specifically, understanding which components of the intervention contributed to observed impact (41); iii) they help to make effectiveness results more interpretable and to avoid type III error, i.e. evaluating an intervention that has not been adequately implemented and therefore drawing incorrect conclusions about an intervention’s effectiveness (42); and iv) they are tools to assess the quality and accuracy of the intervention delivered to participants. This trial’s implementation evaluation will capture information relating to facilitators and barriers for participation in and delivery of LEAPS, as well as fidelity to the LEAPS program model and the quality of the ECCE and youth vocational training services delivered. In doing so, we seek to better understand the processes and conditions required to support program expansion, replication, and sustainable scale-up. *Cost evaluation:* Attention to cost-effectiveness of interventions is a crucial component of intervention planning and evaluation, particularly in resource-limited contexts (41). The LEAPS cost evaluation will provide information about costs to government providers to inform strategies for program expansion and replication. This evaluation will apply an ingredients approach to assessing program costs, cost-per-beneficiary and cost-per-impact (43).

#### Systems strengthening evaluation

This component of the study aims to evaluate the readiness of NCHD to uptake LEAPS for sustainable scaling and replication for expansion in Sindh and other provinces in Pakistan. Our research team will be facilitating a workshop series for NCHD to provide technical capacity building around organizational capacities identified as central to sustainable, quality implementation of LEAPS. Six “pillars” for systems strengthening were identified based on a review of frameworks for supporting, monitoring and evaluating organizational capacity development (44-47), transition readiness (48-50) and community-based management training (51, 52), and informed by inputs and feedback from the NCHD provided during a needs assessment conducted by the research team. The LEAPS System Strengthening Pillars include (i) training, (ii) supervision, (iii) monitoring and evaluation, (iv) leadership and governance, (v) advocacy and communication, and (vi) financing and resource mobilization. A set of indicators will be co-created with NCHD and used to assess NCHD’s “systems readiness” across each pillar. The systems strengthening evaluation has three aims: i) to identify barriers and enablers for implementation at-scale; ii) to assess the effectiveness of the capacity development strategy on NCHD’s competencies related to the LEAPS System Strengthening Pillars; and iii) to evaluate efforts of advocacy in NCHD with partners for replication, expansion, and investment (e.g., awareness about ECCE and youth development, enactment of policies, funding commitments made, sustainability plan developed).

### Outcomes, mediators, and covariates

The trial’s primary (child) and secondary outcomes (youth) and covariates are described in Table 3. All measures will be adapted and piloted for the local socio-cultural and language context (53). The classroom observation tool will be adapted for use in LEAPS from the Measure of Early Learning Environments (MELE) module of the Measuring Early Learning Quality and Outcomes (MELQO) tool (54) and the Instrument for Measuring Quality of Early Childhood Education in Colombia (IMCEIC) (55). Targeted outcomes and covariates are selected to align with the LEAPS program theory of change (see Figure 4).

**Figure 4.**
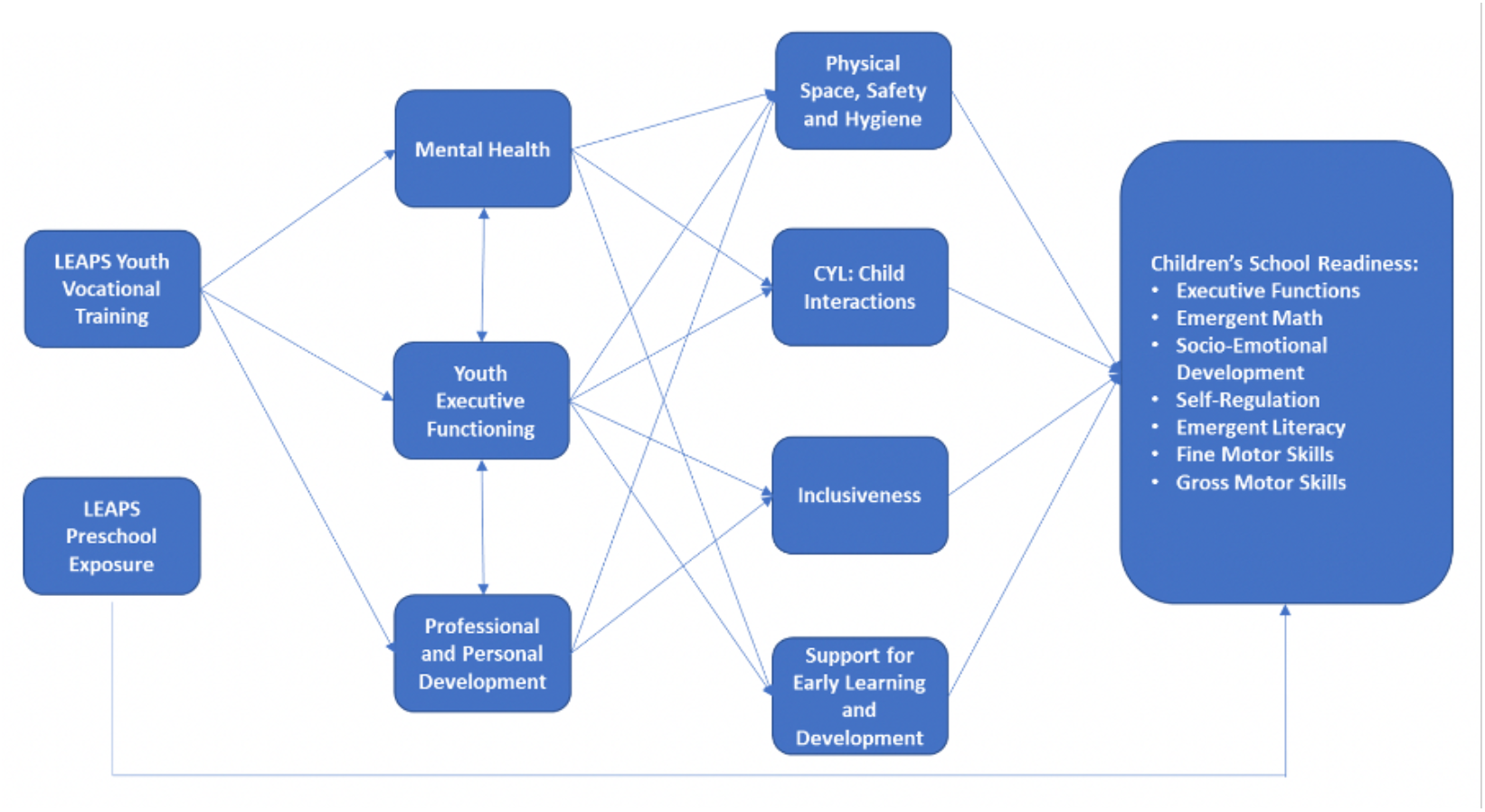
LEAPS Program Theory of Change

### Sample size and power calculation for stepped-wedge cluster randomized trial

A power calculation was conducted with respect to the changes in average child outcomes using representative population surveys. The primary study outcome is children’s school readiness as indexed by scores on the International Development and Early Learning Assessment (IDELA). Based on the results of the LEAPS efficacy trial (26), with a fixed number of clusters (n=99), an intraclass correlation coefficient (ICC) of 0.175, a Step-size of three (n=33 clusters randomized per Step), we estimate that at least 8 children per cluster per round, or 792 children per round, will need to be enrolled to detect a 0.3 SD effect size at 80% power. To achieve adequate power and take refusal rate into account, we will enroll 11 child-caregiver dyads per cluster for a total of 1,089 child-caregiver dyads per round, which should detect a 0.3 SD effect size at 90% power with full recruitment. In other words, we estimated sufficient sample sizes to achieve at least 80% power with up to 30% refusal/under-recruitment rate in each round of the data collection under the intent-to-treat principle. This includes refusals from caregivers and children, or under-recruitment due to decrease in children available at the point of data collection, especially in smaller villages.

### Blinding

While the intent is for the evaluation team to be blinded to intervention status, there are limitations to the extent that assessors can be blinded to participants’ intervention status, given that assessors live locally and intervention activities are visible in communities. However, team members are taking measures to support blinding where possible, including having separate intervention and evaluation field team members and holding separate team trainings and meetings.

### Data collection methods

#### Data collection team

All data collection will be conducted by trained assessors, most of whom have experience working in prior early childhood development trials. Assessors have at least a bachelor’s-level education. Data collection will be overseen by the evaluation field lead and field research manager. All trainings will be conducted by the evaluation field lead and field research manager. Assessors will receive an initial 10-day training for all child and youth assessments, followed by a one-day refresher training prior to round of data collection. To complete training, assessors must have >95% agreement for all child and youth direct assessments with the evaluation field lead. Data collectors engaged in conducting qualitative data collection will receive an additional one-week training in qualitative methods, followed by continued refreshers and debriefing sessions for each qualitative data collection activity. For the classroom observation tool, the field research manager and project coordinator will complete a training-of-trainers with an expert at the Universidad de los Andes, Bogota, Colombia. The field research manager will then train the evaluation field lead with support from the project coordinator. The field research manager and evaluation field lead will then deliver a one-week training to a team of four assessors. Assessors will be required to achieve an interrater reliability of > 70% agreement with a Master Code in coding three 30– 45-minute videos in order to complete training. Coordination meetings will be held daily, with intensive debriefing meetings including capacity building activities held once per week. The evaluation field lead will conduct regular field visits to observe data collection to maintain quality. In each of the three Steps, two assessments per assessor will be observed and independently scored for interrater reliability. Assessments will be video recorded and reviewed with team members during weekly debriefing meetings.

#### Data collection procedures for child and caregiver assessments

Assessors will conduct household visits to issue invitations to the study. If accepted, the assessor will enroll the caregiver in the study on site and administer the household questionnaire to the caregiver during the home visit. The assessor will evaluate the child at the household, and the child will be provided with a rest break and refreshments as needed. Questionnaires will be completed with caregivers using tablets. Data collection for the direct assessments will be conducted manually and then later entered in tablets. At the end of the session the child/caregiver will be given a thank you gift (e.g., book or toy).

#### Data collection procedures for youth participant assessments

CYLs will be issued an invitation to the study during the recruitment and training process. Assessors will issue invitations to comparison youth after completing the screening tool. The youth participant will either be enrolled and assessed on site or at a follow-up appointment, as is practical for the participant. The assessment will be conducted either at a recruitment workshop, training session, or during a home visit with the youth participant. All consecutive follow-up assessments will be conducted at the participant’s household, at the LEAPS school, or at an alternative community space identified for assessments. Location of assessments will be selected based on convenience for the youth participant and the availability of private space. All questionnaires will be collected using tablets. Executive functioning assessments will be conducted manually and later entered in tablets. Youth participants will receive a thank you gift (e.g., a LEAPS mug) and refreshments for participation.

#### Data collection procedures for classroom observations

Assessors will first obtain permission from the local government and school leadership for the observations, after which they will visit schools to coordinate with the teacher and supervisor to schedule the observation. Assessors will arrive before the start of the school day when possible. Assessors should ensure that they are able to observe a minimum of 2.5 hours of the classroom session. If there is less than 2.5 hours of the session remaining, the observation should be rescheduled. If a teacher/CYL is absent, the observation will be rescheduled. Assessors should conduct a maximum of two visits: if the teacher/CYL is absent on the first visit, the assessor will reschedule and conduct a second visit. If the teacher/CYL is absent on the second visit, then the school will no longer be included in the data collection round. The observation may be video recorded as an additional resource to reference when determining final scores. Assessors will observe the class without interrupting the school routine or activities. Assessors will take notes throughout the session and manually complete the observation tool after the session. Videos may be reviewed for items where there is a discrepancy in scoring.

#### Qualitative data collection procedures

All IDIs and FGDs will be conducted by data collectors trained in interview techniques. All activities will be conducted to ensure the participant’s privacy and comfort, and to minimize risk of disturbances. The IDIs and FGDs will be semi-structured, using pre-piloted topic guides and will be recorded. The FGDs will additionally be observed by a data collector trained as a note-taker.

### Data management

Data is collected on encrypted tablets and will be uploaded from the tablets to an encrypted server maintained by the Aga Khan University Data Management Unit (DMU). Paper-based assessments (i.e., classroom observation tool) will be entered in the DMU database using FoxPro by the DMU research assistant at the field office. Initial checks for quality assurance will be conducted by the DMU research assistant and a DMU data coordinator in collaboration with the evaluation field lead. Data will be shared via regular secure transfers with the team members at Harvard, where it will be uploaded to a password-protected shared Dropbox folder. Access to the data folder will only be given to the research team. At Harvard the database will be managed by the data analyst and project coordinator, with regular cleaning and checks for quality assurance. All data will be de-identified and the keys linking identifiers to participants will be stored in a password protected folder. Data will be wiped from the tablets after the round of data collection activities have been completed. All video and audio recordings will be securely stored on a password-protected computer at the field office managed by the evaluation field lead and will be deleted from the camera/recorder after uploading. Recordings will be deleted after analysis is complete (i.e., within one year of endline). Paper-based data assessments will be stored in a locked cabinet at the field site.

Qualitative data will be transcribed in Sindhi by assessors. Five percent of recordings will be randomly chosen by the field research manager and evaluation field lead to check quality of transcription. Transcripts will be translated from Sindhi to English by a translator outside of the project team. Five percent of translations will be chosen randomly for independent back translations. Transcripts, notes, and photos from original qualitative data collection activities will be managed through a password-protected shared Dropbox folder. Only research team members will have access. All data will be de-identified and the keys linking identifiers to participants will be stored in a password protected folder. Audio recordings will be securely stored on a password-protected computer at the field office managed by the evaluation field lead and will be destroyed after the project has been completed.

### Analysis

#### Impact evaluation

The primary analysis will use the intent-to-treat principle. Baseline balance will be tested between randomized groups to see whether randomization was successful. The primary outcome, continuous IDELA score, will be calculated for each individual. We will assess the effect of the LEAPS intervention using the analytical framework proposed by Hussey and Hughes (71) for analysis of stepped wedge cluster randomized trials. Estimation of treatment effects will be obtained from a generalized linear mixed model (GLMM) to produce the mean difference in IDELA scores for LEAPS intervention versus control. The GLMM will include a fixed effect for stratified randomization group, a random effect for cluster, a fixed effect for time, and use an exchangeable correlation structure. In addition, the study will perform a set of sensitivity analyses to explore potential deviations from the basic SW-CRT model proposed by Hussy and Hughes (71). The closed school cohort data will be to examine the changes in the average school readiness among children participating in the LEAPS preschool between baseline and endline. The preschool classroom structural and process quality data will serve as a mediator in the mediation analyses, using the structural equation modelling technique.

#### Implementation and systems readiness evaluations

Secondary administrative data for program monitoring will be analyzed using Excel and Stata to examine trends in program quality indicators over time and by subgroups (e.g., district, FO, CYL). Translated transcripts of the semi-structured IDIs and FGDs will be analyzed through a thematic analysis. Team members will independently review transcripts, followed by a combination of inductive and deductive coding and discussion to identify themes. A codebook and coding matrices will be constructed to support coding and analysis. Triangulation between stakeholders and data sources will be analysed to understand both converging patterns and unique perspectives.

#### Cost evaluation

A cost analysis will be conducted to estimate total project costs for all stakeholders, in addition to cost per beneficiary and cost per impact. Costing data will inform NCHD efforts for expansion and replication of LEAPS.

## Ethics and dissemination

### Protocol amendments

All modifications to the protocol will be communicated with the PI, co-PI and co-investigators through regular meetings and email communications. Any amendments to the protocol that have implications for the study design or participant safety will be reported to the affiliated ethics review boards through applications and email communications and to the trial registry through the application process. Protocol amendments impacting trial participants will be communicated in-person by the field research manager, evaluation field lead, or trained assessors.

### Reporting and risk of adverse events and harms

The LEAPS research team will be responsible for monitoring regulatory documents and participant files, including reviewing and maintaining all required ERC/IRB documentation, monitoring and updating regulatory supporting documents, recording meeting minutes, documenting the recruitment, enrollment, and informed consent processes, and maintaining logs to track data collection and assessment activities. There is a small risk of loss of confidentiality for participants. To minimize this risk, interviews will be conducted in private spaces; tablets will be password protected; hard copies of forms will be stored in locked cabinets; data will be de-identified prior to sharing; and assessors will receive training in good clinical practice.

There is additionally a risk of psychological discomfort for youth participants due to survey questions on depressive symptoms. There is minimal risk associated with administration of the child assessments for executive function skills and school readiness and the household questionnaire with caregivers. To mitigate these risks, assessors are trained in how to sensitively administer youth and child assessments, and how to communicate with children’s parents about the assessment and their child’s performance. Participants will be informed that they may stop at any time without consequence.

The youth participant questionnaire includes a question on suicidal ideation. If the participant responds positively to this item, the assessor will contact the evaluation field lead and field research manager who will assess whether there is an imminent danger of self-harm. If there is immediate danger, then the assessor will escort the participant to a health provider. The team will also discuss with a consultant at the Aga Khan University Department of Psychiatry as needed. In no instance will the person be left alone. If the person has suicidal ideation, but is not at risk of immediate self-harm, the study staff will make an active referral for mental health care.

Finally, corporal punishment is unfortunately widely accepted in the intervention communities, so there is a risk that CYLs may initially use harsh disciplinary practices in the LEAPS classrooms. To mitigate this risk, CYL training includes preschool behavior management, covering strategies for appropriate classroom management and employing gentle discipline. The NCHD’s CYL contracts also include a clause prohibiting use of corporal punishment. If a CYL is observed using harsh discipline in the classroom, she will receive targeted training in classroom management. Repeated episodes of harsh discipline by a CYL will result in termination of the CYL’s employment.

### Consent to participate

For all data collection activities, informed consent will be obtained from participants by the research team’s trained assessors. For participants under age 18 years, both child assent and consent from a legal guardian will be obtained. Consent forms were written to explain the information with accuracy and clarity for individuals with low levels of literacy (on average mothers in this location have two years of formal education) (26, 56). All forms are written in Sindhi, the local language. All subjects will be free to withdraw from the study at any time and will be assured that this will not affect the standard of care received in the community.

Consent for the preschool classroom observations and video recording will be obtained from the CYL/comparison preschool teacher as well as a representative from the school’s parent-community board. For schools that do not have a parent-community board, consent will instead be sought from the school’s leadership, i.e., the school principal or coordinator. Consent will be required from both the CYL/comparison preschool teacher and the representative from the school board or school leadership in order to proceed with the video recordings. The assessor will then enroll the CYL/teacher at the school and observe the classroom session to complete the assessment.

### Confidentiality

All assessments will be conducted in a private location, either in an assessment center, classroom, office, or private household. All data (including recorded interviews and notes, direct assessments, questionnaires, video recordings for quality assurance) will be de-identified by the research team. The keys linking identifiers to participants will be stored in a password protected file (separated from the other data files), to which only the research team members will have access. All hard copy materials containing identifiable information (i.e., consent forms, interview cover sheets, photos) will be stored in locked cabinets. All tablets used for data entry will be password protected. Tablets and video recorders will be stored in locked cabinets when not in use. Data will be regularly transferred to the central server at Aga Khan University and then wiped. Video and audio recordings will be deleted after analysis is completed (within one year of endline). All research team staff will receive training in protecting participant privacy and adherence to Good Clinical Practices.

### Ancillary and post-trial care

Given that LEAPS serves vulnerable populations with limited access to education, training, and employment, the research team has a responsibility to ensure that LEAPS participants are supported beyond the trial period. The research team will coordinate with NCHD to ensure that, a) LEAPS children have a clear pathway to transition to primary school at the close of the trial, and b) that CYLs are supported through career counseling exit meetings to identify further options for education and employment.

### Dissemination: trial results

Results will be disseminated through a variety of channels, including workshops with government stakeholders, funder and lay reports, presentations at national and international conferences, publications in peer-reviewed journals, and through networks for early childhood and youth development such as Asia-Pacific Regional Network for Early Childhood, the Pakistan Alliance for Early Childhood, the Early Childhood Development Action Network, Early Childhood Peace Consortium, and the Alliance for International Youth Development.

## Discussion

This will be the first larger-scale impact evaluation of a youth-led ECCE program in an LMIC, providing much-needed empirical support for the youth-led ECCE model. The results of this evaluation will enable Pakistan’s NCHD to make evidence-based decisions about strengthening and expanding their community-based preschool and youth vocational training systems. The evidence from this evaluation will also be of relevance to regional and global settings where there is a vast unmet need for preschool services and limited employment and learning opportunities for youth. This study will additionally contribute to the literature around capacity development to support sustainable transitions of programming from donors to government ownership with changing levels of support technical partners, providing evidence from a systematic approach to capacity building and program transition with government partners in a rural, resource-limited context. It is our vision that this trial will lay the groundwork for future efforts to expand and replicate LEAPS in other NCHD communities, in order to strengthen NCHD’s ECCE programming on a wider, systems-level. Figure 5 depicts the end goal for a fully integrated LEAPS program model, linked to NCHD Feeder Primary Schools within the Universal Primary Education Program. An expanded program model would additionally aim to obtain accreditation for CYL vocational training. Finally, findings from this work will contribute to the research around gender equity and youth development, providing a rigorous evaluation of a female-specific youth development program in an LMIC, a context for which there are very few program evaluations (18).

**Figure 5.**
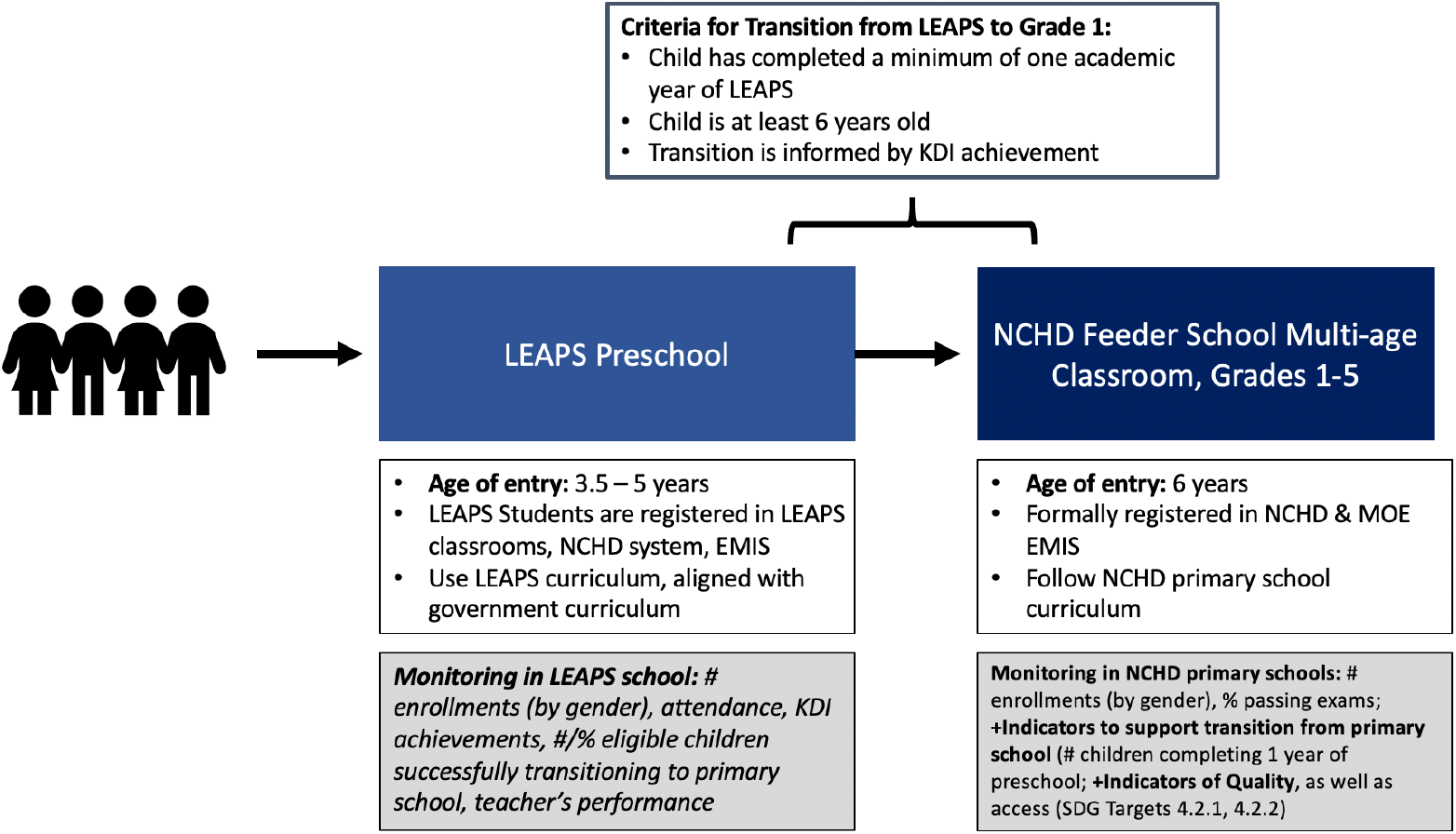
NCHD Universal Primary Education Program (UPE) Feeder School and LEAPS Programs, Under Scale-up. This diagram presents the vision for the NCHD Universal Primary Education program structure including the Feeder schools and LEAPS schools under scale-up; KDI = Key Development Indicator; NCHD = National Commission for Human Development; SDG = Sustainable Development Goals.

## Limitations

A limitation of the stepped-wedge design is that time is associated with both the exposure to the intervention and potentially the outcome, and in this way is a possible confounder (40). Calendar time will therefore be adjusted for in the analysis. Potential interruptions due to unexpected emergency contexts, such as emergency shutdowns and flooding, also pose a risk to the study design and implementation. Finally, because the project is implemented in collaboration with a government organization, turnover in government partners may create challenges for program delivery.

## Trial Status

This trial is currently active (protocol version 5, April 9, 2021; see Appendix 2). Recruitment of participants began in January 2019. Final participants are expected to be recruited and complete their assessments at the end of May 2021.

## Data Availability

Following publication of all planned manuscripts, the final de-identified dataset will be stored indefinitely on the Harvard Open Data Dataverse
Network, except for in the case of request of withdrawal by the participant for themselves or the child in which case the data will be deleted.
Permission to use the data should be sought from the PI in advance, and the source should be cited.

## Abbreviations

AKU: Aga Khan University
CYL: Community Youth Leader
DMU: Data Management Unit
ECD: Early Childhood Development
ECCE: Early Childhood Care and Education
FGD: Focus Group Discussion
FO: NCHD Field Officer
HFIAS: Household Food Insecurity Access Scale
IDELA: International Development and Early Learning Assessment
ICC: intraclass correlation coefficient
IDI: In-depth interview
IMCEIC: Instrument for Measuring Quality of Early Childhood Education in Colombia
KDI: Key Development Indicator
LEAPS: Youth Leaders for Early Childhood Assuring Children are Prepared for School
LHW: Lady Health Worker
LIST: LEAPS Intervention Support Team
LMIC: Low- and Middle-Income Country
MELE: Measure of Early Learning Environments
MELQO: Measuring Early Learning Quality and Outcomes
NCHD: National Commission for Human Development
PI: Principal Investigator
PTM: Parent Teacher Meeting
PYD: Positive Youth Development
SDG: Sustainable Development Goal
SPIRIT: Standard Protocol Items: Recommendations for Interventional Trials
SRQ-20: Self-Reporting Questionnaire 20-Item
TiDiERS: Template for Intervention Description and Replication
TMT: Trail Making Task
UPE: NCHD Universal Primary Education program

## Declarations

## Research ethics approval and consent to participate

All protocol, consent forms, and data collection tools were reviewed and approved by the Institutional Review Board of the Harvard T. H. Chan School of Public Health, Boston, MA, USA (Protocol #: IRB18-1149); the Ethics Review Committee at Aga Khan University Karachi, Pakistan (5473-Ped-ERC-18); and the National Bioethics Committee Pakistan, Islamabad, Pakistan (Ref: No.4-87/NBC-334-Y2-Extension/19/). All participants/legal guardians will have the provided informed consent to participate.

## Consent for publication

Not applicable.

## Availability of data and materials

Following publication of all planned manuscripts, the final de-identified dataset will be stored indefinitely on the Harvard Open Data Dataverse Network, except for in the case of request of withdrawal by the participant for themselves or the child in which case the data will be deleted. Permission to use the data should be sought from the PI in advance, and the source should be cited.

## Competing interests

The authors have no competing interests.

## Funding

Funding for this work is provided by Dubai Cares and Saving Brains Program Grand Challenges Canada (award number TTS-1808-17846). The funder has no role in the development or implementation of the study, nor will they be involved in the analysis, interpretation, report writing, or publication and dissemination.

## Authors’ contributions

AKY conceptualized the study, led the development of the LEAPS intervention, identified outcomes measures, contributed to trial design and plan of analysis. CS designed the study trial and contributed to the plans for analyses. EF and SS adapted measures for outcome data collection for the study. EF contributed to the plan of analyses. SS, EF, KR developed the intervention components of the LEAPS Programme and qualitative topic guides. CR, LP contributed to the development of the intervention and quantitative and qualitative measures. SB contributed to the qualitative topic guides and to the development of the intervention. GF contributed to the study design, measures and plans for analyses. RD contributed to the plan of analysis. AKY, EF, KR, RD prepared the first draft. All authors reviewed, provided critical feedback and approved the final protocol.

## Acknowledgements

We sincerely thank the National Commission for Human Development Head Office in Islamabad, Province Office in Karachi, Sindh, and four District Offices in Sindh province, including district Dadu, Khairpur, Naushahro Feroze, and Sukkur, for their invaluable partnership and support in carrying out this study, as well as the LEAPS families, children and CYLs for their support and commitment. We are deeply grateful to Ms. Shahnaz Parveen Hakro and Tasleem Soomro for their dedication and leadership. Finally, we thank Mr. Amjad Hussain, Mr. Zubair Tagar, and Mr. Hassan Naqvi with the Aga Khan University Data Management Unit for their support in evaluation and data collection efforts.

## Appendix 1.

Supplemental tables and figures

**Table 1.** Child and parental education in rural Sindh, 2019. Summary of school enrollment, completion, and learning outcomes for children and their caregivers in rural communities of Sindh Province, Pakistan (ASER Pakistan, 2020).

| Indicator | Outcomes in rural Sindh |
| --- | --- |
| **Preschool Enrollment (aged 3-5 years)** |  |
| Overall | 45.6% |
| By Age Group |  |
| 3 years | 21.0% |
| 4 years | 67.2% |
| 5 years | 23.4% |
| **Basic Education Enrollment (aged 6-16 years)** |  |
| Overall | 86.2% |
| By Age Group |  |
| 6-10 years | 84.7% |
| 11-13 years | 92.2% |
| 14-16 years | 84.6% |
| By Gender |  |
| Boys | 62.4% |
| Girls | 37.6% |
| **Child Learning Outcomes (aged 5-16 years)** |  |
| *Learning Levels, Sindhi (5-16 years)* |  |
| Grade 1: Can read letters | 44.4% |
| Grade 3: Can read sentences | 23.5% |
| Grade 5: Can read stories | 24.0% |
| Can read at least sentences: |  |
| Boys | 33.0% |
| Girls | 25.0% |
| *Learning Levels, English (6-16 years)* |  |
| Grade 1: Can read letters | 38.8% |
| Grade 3: Can read words | 26.9% |
| Grade 5: Can read sentences | 26.6% |
| Can read at least words: |  |
| Boys | 32.0% |
| Girls | 24.0% |
| *Learning Levels, Arithmetic (6-16 years)* |  |
| Grade 1: Can recognize numbers 1-99 | 48.2% |
| Grade 3: Can do two-digit subtraction | 21.7% |
| Grade 5: Can do two-digit division | 30.5% |
| Can at least do subtraction: |  |
| Boys | 28% |
| Girls | 22% |
| **Parental Education (among parents with children aged 6-16 years)** |  |
| Completed Primary School: |  |
| Mothers | 26% |
| Fathers | 44% |

**Table 2.** Youth education & employment in rural Sindh and rural Pakistan, 2018-2019. Summary of literacy rates among male and female youth aged 15-24 years in rural Sindh, top reasons for leaving school early among male and female children and youth aged 10-18 years in rural Sindh, school attendance and school completion among male and female youth aged 15-24 years in rural Pakistan, and national workforce participant rates and unemployment rates among male and female youth aged 15-24 years (Pakistan Bureau of Statistics, 2018; Pakistan Bureau of Statistics, 2020).

|  | Male |  | Female |  |
| --- | --- | --- | --- | --- |
| Rural Sindh |  |  |  |  |
| Youth Literacy (among 15-24-year-olds) | 61% |  | 30% |  |
| Top Reasons for Leaving School Early (among 10-18-year-olds) | Student not willing (50%) |  | Student not willing (30%) |  |
|  | Had to help at work (10%) |  | Parents didn't allow (16%) |  |
|  | Other (27%) |  | Other (24%) |  |
|  | Aged 15-19 years | Aged 20-24 years | Aged 15-19 years | Aged 20-24 years |
| Rural Pakistan |  |  |  |  |
| Ever Attended School | 78.0% | 59.0% | 74.0% | 52.0% |
| Completed Primary School | 72.0% | 69.0% | 54.0% | 48.0% |
| Nationally in Pakistan |  |  |  |  |
| Labor Force Participation Rate | 47.6% | 84.6% | 15.6% | 23.3% |
| Unemployment Rate | 11.6% | 10.5% | 6.5% | 15.2% |

**Table 3.**
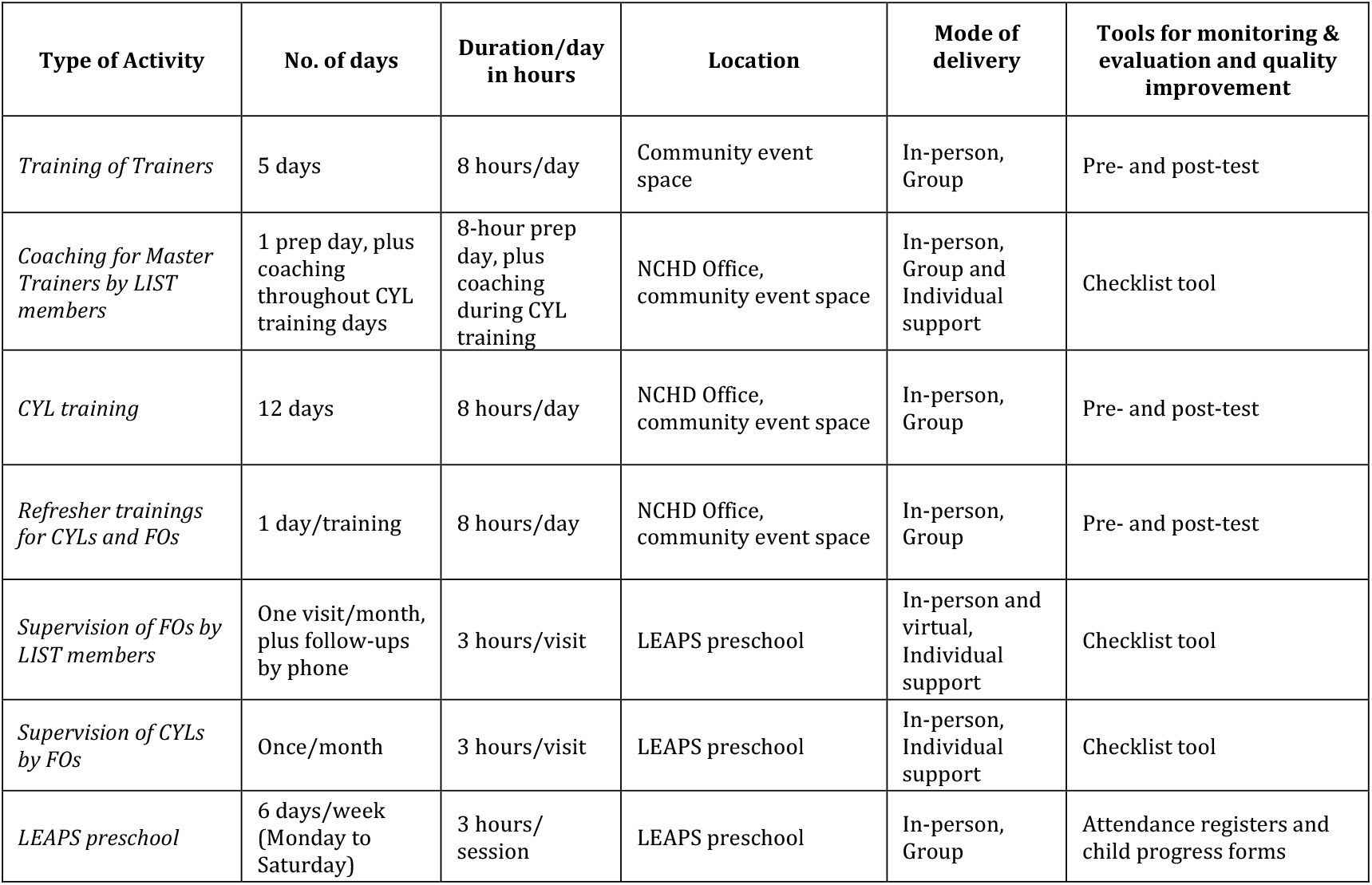
Summary of LEAPS intervention delivery. This table outlines the intervention dosage, location, modes of delivery, and tools for monitoring and evaluation for the LEAPS program. CYL= Community Youth Leader; FO= Field Officer; LIST = LEAPS Intervention Support TEAM; NCHD = National Commission for Human Development.

**Table 4.** Overview of LEAPS CYL Training. Summarizes the 12-day CYL training in detail, including the duration, format, training content, and manuals and forms given to CYLs; CYL=Community Youth Leader; NCHD = National Commission for Human Development; LIST = LEAPS Intervention Support Team.

| Duration and Format of Training | Training Content | Manuals and forms given to CYLs |
| --- | --- | --- |
| The 12-day teacher training is held at each of the four districts at either NCHD office or other community space. Each training group has a maximum of 15 CYL participants. These trainings are eight hours in duration each day.<br>The first 10 days include icebreakers, discussion, and role-play exercises. The remaining two days are scheduled for "hands-on" practice or practicum opportunities. | During the first 10 days, CYLs will review early childhood development modules and LEAPS preschool curriculum in detail. Trainers follow a structured training manual.<br>During the remaining two days, CYLs will be offered opportunities to practice activities from the preschool curriculum. These practicums are held at an already running LEAPS preschool, with support from NCHD Field Officers and LIST members. | • **Community-Based Preschool Activity guide:** Details classroom-based activities which are categorized according to 50 Key Development indicators (KDIs) across the seven LEAPS learning areas. These seven learning areas include: (i) Approaches to learning; (ii) Social-emotional, and moral development; (iii) Physical development and health; (iv) Language, literacy, and communications; (v) Mathematics; and (vi) My world.<br>• **Attendance and Enrolment Registers:** A session focusing on filling attendance and other contact information is conducted to familiarize CYLs with how to use their registers.<br>• **Child Progress Forms:** A copy of child progress forms will be provided to CYLs. CYLs will use this form to assess children's performance and progress toward achieving the KDIs throughout the academic year. |

**Table 5.** LEAPS Preschool Routine. Table 5 describes the LEAPS preschool routine and activities; CYL = Community Youth Leader.

| Time | Session | Routine |
| --- | --- | --- |
| 8:00- 8:30 am | Arrival Time and Free Play | • Meet and greet children and parents<br>• Help children wash hands<br>• Children self-register attendance on the attendance chart<br>• Free Play: Children can play in groups or individually in the quiet corner designated in each classroom |
| 8:30- 8:45 am | Snack Time | • Wash hands<br>• Children take their snack<br>• Children clean up after snack with support from CYL |
| 8:45- 9:10 am | Circle Time | • CYLs share stories, songs, rhymes, etc. during circle time. Additionally, they use circle time to talk about the day of the week, weather, and any other special information (e.g., a child's birthday) |
| 9:10- 9:20 am | Break | • 10-minute stretch break |
| 9:20-10:05am | Learning centers | • Individual and small group activities |
| 10:05-10:10am | Clean-up Time | • CYL sings clean up songs with children and supports them to clean up |
| 10:10-10:50 am | Large group activities | • Children play indoors or outdoors in large group (e.g., action songs or playing with a ball). |
| 10:50-11:00 am | Departure Time | • CYL reviews the day with children<br>• Helps children to wash hands and say goodbye |

**Table 6.** LEAPS Actions for Emergency Response: Table 6 shows three response actions for LEAPS that will be implemented during times of emergencies or conflict in which school routines are disrupted. CYL= Community Youth Leader; KDIs= Key Development Indicators.

| Response Action | Descriptions |
| --- | --- |
| Remote Learning Activities for CYLs | CYLs may engage in remote learning activities ranging from writing a story to creating learning materials for their classroom. Remote-learning activities are intended to help CYLs to stay connected to LEAPS and support skill development. |
| Workbooks for Children | In order to promote learning at home and to stay connected to LEAPS, children will be given a workbook during times of emergencies or conflict. The children's workbook will have several learning worksheets which children can complete with support from their family members and siblings. Example worksheet topics will include alphabets, numbers, transportation, local animals, fruits, and vegetables. These worksheets will be aligned with the 50 KDIs across seven learning areas. |
| Additional Needs-Based Trainings | Additional trainings will be conducted prior to school re-opening especially if schools have been shut down for a longer period of time. These trainings will help CYLs review concepts from the initial 12-day training and also address additional content arising from emergency and conflict situations which may not have been previously addressed. Where needed, additional resources such as handouts, storybooks, classroom materials, etc., will be provided to support CYLs and children. |

## Appendix 2.

Supplemental tables and figures

### Section 1. Protocol Version

**Issue Date:** April 9, 2021

**Protocol Amendment Number:** 04

**Authors:** AKY, SB

**Table 1.** Revision Chronology

|  |  |
| --- | --- |
| **December 5, 2018** | Original |
| **January 25, 2019** | Amendment 01: Primary reason for amendment:<br>Updated Study status to 'recruiting'. |
| **June 6, 2019** | Amendment 02: Primary reason for amendment:<br>Updates to study contacts |
| **July 31, 2020** | Amendment 03: Primary reason for amendment:<br>Extension of study period due to COVID-19 interruptions. Updates to IDELA scoring information. |
| **April 9, 2021** | Amendment 04: Primary reason for amendment:<br>Updated outcome measures to specify child executive functioning outcomes as a secondary outcome measure in order to limit the study to a single primary outcome measure of school readiness indexed by IDELA (determined a priori). |

### Section 2. Sponsor Contact Information

**Trial Sponsor:** Harvard

**Sponsor’s Reference:**

***Contact Name:*** Aisha K. Yousafzai

***Address:*** 665 Huntington Ave., Boston, MA 02115

***Telephone***: +1-857-318-8691

***Email***

## Notes

### Competing Interest Statement

The authors have declared no competing interest.

### Clinical Trial

NCT03764436

### Author Declarations

All protocol, consent forms, and data collection tools were reviewed and approved by the Institutional Review Board of the Harvard T. H. Chan School of Public Health, Boston, MA, USA (Protocol #: IRB18-1149); the Ethics Review Committee at Aga Khan University Karachi, Pakistan (5473-Ped-ERC-18); and the National Bioethics Committee Pakistan, Islamabad, Pakistan (Ref: No.4-87/NBC-334-Y2-Extension/19/).

